# Deep Learning-Enabled Screening of Chronic Kidney Disease from Echocardiography

**DOI:** 10.64898/2026.02.02.26345379

**Authors:** Victoria Yuan, Hirotaka Ieki, Alexander Sandhu, Long H. Nguyen, Paul P. Cheng, Stephanie T. Chang, Andrew P. Ambrosy, Alan C. Kwan, Alan S. Go, Susan Cheng, David Ouyang

## Abstract

Chronic kidney disease (CKD) affects nearly 850 million individuals globally; the prevalence of undiagnosed CKD is 60%. Taking advantage of the relationship between CKD and cardiovascular disease, we developed a deep learning (DL) model to detect CKD from parasternal long-axis (PLAX) videos using 325,377 PLAX videos from 62,818 patients at Cedars-Sinai Medical Center (CSMC). We externally validated our model in two independent cohorts of 2,224 patients at Stanford Healthcare (SHC) and 41,611 patients at Kaiser-Permanente Northern California (KPNC). In a held-out test cohort at CSMC, our model detected any stage of CKD with an area under the curve (AUC) of 0.756 [95% confidence interval 0.749 – 0.763], with consistently strong performance in KPNC (AUC 0.718 [0.714 – 0.723]) and SHC (AUC 0.719 [0.704 – 0.735]). Our DL echo model detected CKD with robust performance at two external clinical sites, offering an avenue for noninvasive screening and improved detection rates.

## Introduction

Nearly 850 million individuals worldwide are estimated to have chronic kidney disease (CKD)^1^. Because the early stages of CKD may be asymptomatic, the prevalence of undiagnosed CKD ranges from 61.6% to 95.5%^2,3^. Low rates of detection contribute significantly to the growing burden of CKD^1,3–7^. Current screening guidelines require both serum and urine laboratory testing^8^. Although annual testing with urine albumin-to-creatinine ratio (UACR) is standard of care in patients with diabetes mellitus, screening with UACR has rates as low as 20%^9^. Additionally, such screening is rarely performed in patients with other CKD risk factors, such as hypertension or dyslipidemia^6,10^. Suboptimal detection prevents timely management and places patients at risk of disease progression and complications^11^.

CKD and cardiovascular disease (CVD) have a bidirectional relationship and frequently co-occur^7,12–15,16,17^. Patients with CKD face a higher risk of cardiac abnormalities, such as coronary artery disease, left ventricular hypertrophy, and myocardial fibrosis^7,18^. Similarly, heart disease is a risk factor for the development of CKD and contributes to its rising incidence^1,19^. Cardiovascular-kidney-metabolic (CKM) syndrome captures the interactions between CKD, CVD, and their metabolic risk factors, which can lead to excess morbidity^15^. The relationship between CKD and CVD suggests that cardiac imaging may be leveraged to detect renal dysfunction. Early detection of CKD is imperative to connect patients to cardioprotective and renoprotective therapies.

Echocardiography is a widely used, noninvasive imaging modality that is frequently performed to evaluate cardiac pathologies. Given the co-occurrence of CVD and CKD, echocardiography offers an avenue for opportunistic screening of CKD. Machine learning (ML) applied to echocardiography has demonstrated great promise to enhance image evaluation and diagnostics. Current efforts have diagnosed cardiac diseases such as valvular pathologies^20–23^, amyloidosis^24,25^, and cardiomyopathy^26–28^, in addition to extra-cardiac pathologies such as liver disease^29^ and elevated blood urea nitrogen^30^. Given its success in detecting a wide range of pathologies, ML can facilitate identification of CKD from cardiac imaging and address the need for improved detection.

To this end, we present a deep learning model that detects CKD from parasternal long axis (PLAX) echocardiogram videos developed using data from 62,818 patients at Cedars-Sinai Medical Center (CSMC) and externally validated it in a cohort of 2,224 patients at Stanford Healthcare (SHC) and 41,611 patients from Kaiser-Permanente Northern California (KPNC). With consistently strong performance in both cohorts, our model can improve CKD screening and promote the use of deep learning to bridge diagnostic gaps.

## Results

### Study population

Our model was developed using data from 62,818 patients who received care at CSMC and had 146,632 echocardiography studies with 325,377 PLAX videos. The case cohort consisted of 10,472 patients with 24,084 studies and 51,483 PLAX videos who had an ICD-10 code for any stage of CKD and echocardiogram within 1 month of diagnosis. The control cohort included 52,346 patients who had never been diagnosed with an ICD-10 code for CKD and who had 122,547 echocardiography studies with 273,894 PLAX videos. Demographic and clinical information of the case and control cohorts are presented in Table 1, and in the training, validation, and test sets in Supplementary Table 2. The case cohort had higher prevalence of hypertension and diabetes mellitus, which are both known risk factors for CKD. LVEF, LV systolic and diastolic volumes, rate of CAD, and rate of dyslipidemia were similar between case and control cohorts. After splitting the cohort on the patient-level into train, validation, and test sets in an 8:1:1 ratio, the internal test set consisted of 6,282 patients with 14,584 studies and 32,483 PLAX videos.

**Table 1.** Demographics and co-morbidities of the internal development cohort at Cedars-Sinai Medical Center (CSMC) and the external validation cohort at Kaiser-Permanente.

|  | **CSMC**<br>**Controls**<br>**(n=52,346)** | **CSMC**<br>**Cases**<br>**(n=10,472)** | **KPNC**<br>**Controls**<br>**(n=31,357)** | **KPNC**<br>**Cases**<br>**(n=10,254)** | **SHC**<br>**Controls**<br>**(n=1,352)** | **SHC Cases**<br>**(n=404)** |
| --- | --- | --- | --- | --- | --- | --- |
| Age | 67.7 ± 15.6 | 66.5 ± 15.2 | 69.2 ± 12.9 | 73.1 ± 13.1 | 61.0 ± 17.4 | 66.3 ± 14.9 |
| Sex |  |  |  |  |  |  |
| Male | 33,645<br>(64.3%) | 6,731<br>(64.3%) | 15,763<br>(50.3%) | 5,621<br>(54.8%) | 876 (44.6%) | 497 (62.3%) |
| Female | 18,701<br>(35.7%) | 3,741<br>(35.7%) | 15,572<br>(49.7%) | 4,631<br>(45.2%) | 1088 (55.4%) | 301(37.7%) |
| Race and Ethnicity |  |  |  |  |  |  |
| American Indian<br>or Alaska Native | 106 (0.2%) | 28 (0.3%) | 154 (0.5%) | 62 (0.6%) | 7 (0.4%) | 2 (0.3%) |
| Asian | 3,594 (7.0%) | 895 (8.6%) | 4,695<br>(15.0%) | 1,805<br>(17.6%) | 384 (19.6%) | 119 (14.9%) |
| Black or African<br>American | 6,072<br>(11.8%) | 2,238<br>(21.4%) | 2,191 (7.0%) | 1,504<br>(14.7%) | 103 (5.2%) | 54 (6.8%) |
| Native<br>Hawaiian/Pacific<br>Islander | 142 (0.3%) | 46 (0.4%) | 219 (0.7%) | 167 (1.6%) | 32 (1.6%) | 23 (2.9%) |
| Hispanic | 5,738<br>(11.2%) | 1,715<br>(16.4%) | 2,923 (9.3%) | 1,154<br>(11.3%) | 198 (10.1%) | 111 (13.9%) |
| Non-Hispanic | 44,390<br>(86.3%) | 8,652<br>(82.6%) | 28,434<br>(90.7%) | 9,100<br>(88.7%) | 1,635<br>(83.2%) | 672 (84.2%) |
| White | 36,612<br>(71.1%) | 6,484<br>(61.9%) | 17,152<br>(54.7%) | 5,454<br>(53.2%) | 1,021<br>(52.0%) | 437 (54.8%) |
| Other | 3,702 (7.2%) | 664 (6.3%) | 185 (0.5%) | 37 (0.3%) | 309 (15.7%) | 143 (17.9%) |
| Unknown | 2,118 (4.0%) | 98 (0.9%) | 191 (0.6%) | 14 (0.1%) | 108 (5.5%) | 20 (2.5%) |
| Left ventricular<br>ejection fraction % | 52.8% ±<br>17.4% | 56.9% ±<br>14.9% | 57.9% ±<br>10.2% | 53.5% ±<br>13.4% | 57.9% ±<br>11.0% | 51.0% ±<br>15.4% |
| Left ventricular end-diastolic volume index (mL/m<sup>2</sup>) | 46.6 ± 25.8 | 51.9 ± 27.8 | 49.7 ± 17.1 | 55.7 ± 23.8 | 45.8 ± 18.1 | 55.7 ± 30.4 |
| Left ventricular end-systolic volume index (mL/m<sup>2</sup>) | 21.5 ± 17.4 | 25.9 ± 19.7 | 21.7 ± 12.7 | 27.7 ± 20.2 | 20.3 ± 14.1 | 30.3 ± 27.5 |
| Estimated glomerular filtration rate (eGFR, mL/min/1.73m<sup>2</sup>) | 74.02 ± 26.6 | 33.3 ± 22.5 | 58.5 ± 5.7 | 38.1 ± 18.2 | 89.4 ± 21.0 | 57.1 ± 28.5 |
| Urine albumin-to-creatinine ratio (uACR) | 154.3 ± 492.2 | 379.3 ± 737.1 | 46.0 ± 86.4 | 129.2 ± 131.7 | N/A | N/A |
| Coronary artery disease | 12,317 (23.5%) | 4,013 (38.3%) | 13,497 (43.0%) | 6,902 (67.3%) | 407 (20.7%) | 403 (50.5%) |
| Hypertension | 22,572 (43.1%) | 8,334 (79.6%) | 23,436 (74.7%) | 10,009 (97.6%) | 858 (43.7%) | 534 (66.9%) |
| Diabetes mellitus | 9,058 (17.3%) | 5,089 (48.6%) | 9,579 (30.5%) | 7,092 (69.1%) | 477 (24.3%) | 424 (53.1%) |
| Dyslipidemia | 16,739 (32.0%) | 5,171 (49.4%) | 20,857 (66.5%) | 9,021 (88.0%) | 891 (45.4%) | 593 (74.3%) |

The external validation cohort at Kaiser-Permanente had 41,611 patients with 46,970 echocardiography studies and 66,249 PLAX videos. Within this cohort, 10,254 patients with 11,671 echocardiography studies and 16,600 PLAX videos who had been diagnosed with any stage of CKD. 31,357 patients had never received an ICD-10 code for CKD; there were 35,299 studies with 49,649 PLAX videos across these patients. In comparison to the case cohort at CSMC, the case cohort at KPNC had higher rates of diabetes mellitus, hypertension, dyslipidemia, and CAD. The external validation cohort at SHC consisted for 798 case patients with 916 studies and 1,426 PLAX videos and 1, 964 control patients with 2,207 studies and 3,070 PLAX videos. Urine albumin values were not available for the SHC cohort, so UACR was not calculated. Case patients at SHC had a higher mean eGFR and lower rates of diabetes mellitus, hypertension, dyslipidemia, and CAD. Demographics were similar at all three sites.

### Model Performance

Our deep learning model identified any stage of CKD with strong performance in both the CSMC and KPNC cohorts. In the CSMC internal test cohort, our algorithm detected CKD with an AUC of 0.756 [0.749 – 0.763], sensitivity of 75.6% [74.0% - 76.4%], and specificity of 61.3% [61.3% – 62.4%] (Figure 1). The model distinguished CKD with strong performance across the spectrum of disease severity. Performance was consistent across stages of CKD with an AUC of 0.694 [0.670 – 0.718] in discriminating mild CKD, AUC 0.701 [0.689 – 0.712] for moderate CKD, AUC 0.733 [0.755 – 0.790] for severe CKD, and AUC 0.840 [0.829 – 0.850] for ESRD (Supplementary Table 3).

**Figure 1.**
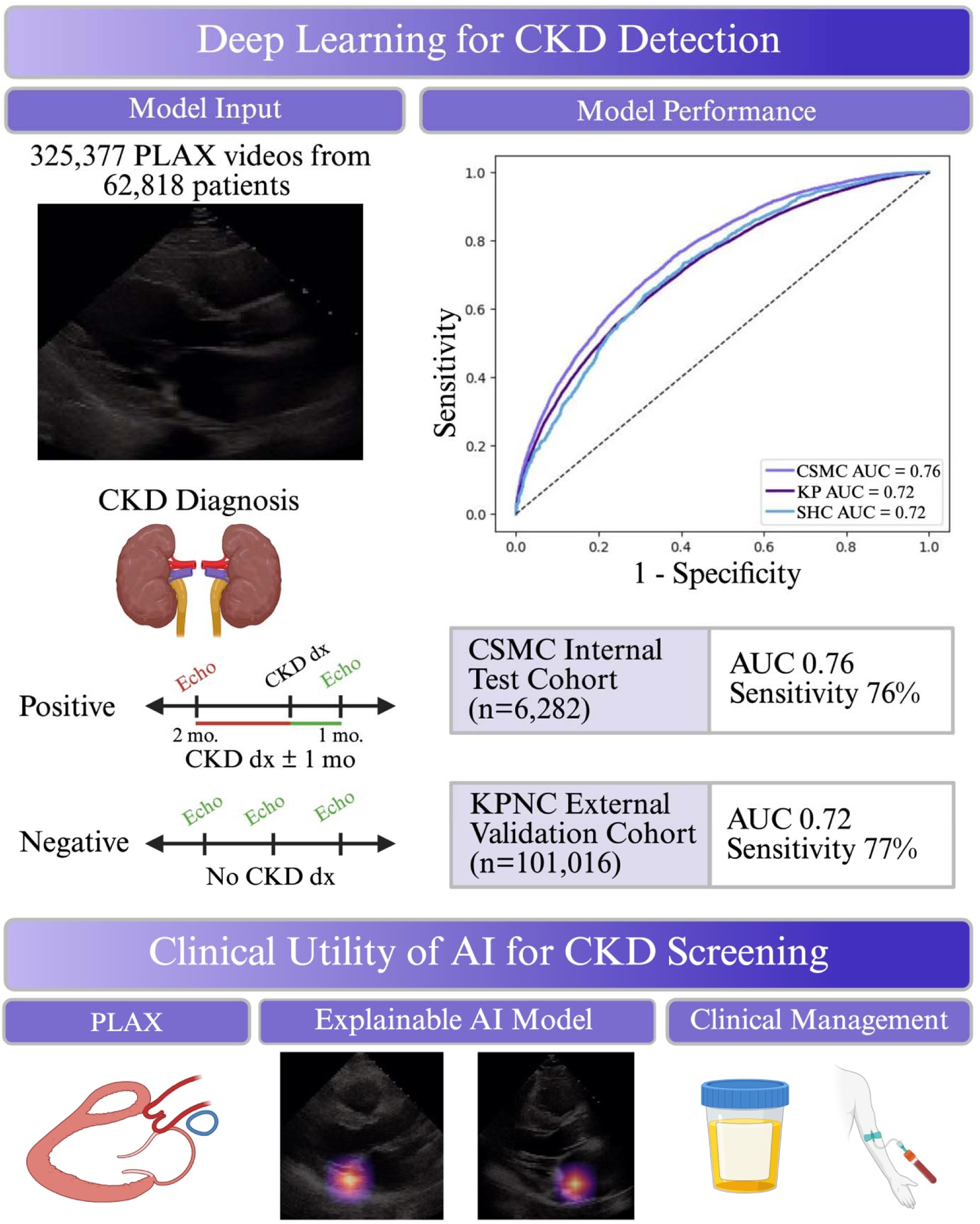
Schematic diagram of model development, model performance in the Cedars-Sinai internal test cohort and Kaiser-Permanente external validation cohort, and the potential utility of interpretable deep learning to screen for CKD.

We then tested model performance in subgroups of patients with diabetes mellitus, hypertension, and hyperlipidemia, as these cohorts are considered higher risk for CKD but are not routinely screened. Our model demonstrated robust performance across subgroups with AUCs of 0.728 [0.716 – 0.739] in patients with diabetes; 0.752 [0.743 – 0.759] in patients with hypertension; 0.756 [0.745 – 0.767] in patients with CAD; and 0.752 [0.742 – 0.761] in patients with dyslipidemia (Table 2, Supplementary Figure 1). Model performance was robust to LVEF (AUC 0.700 [0.685 – 0.715] in patients with LVEF < 50% versus AUC 0.770 [0.762 – 0.779] in patients with LVEF ≥ 50%); sex (AUC 0.762 [0.750 – 0.775] in female patients versus AUC 0.752 [0.743, 0.760] in male patients); and age (AUC 0.820 [0.810 – 0.830] in patients under 65 years old versus AUC 0.708 [0.700, 0.717] in patients equal to or greater than 65 years old).

**Table 2.**
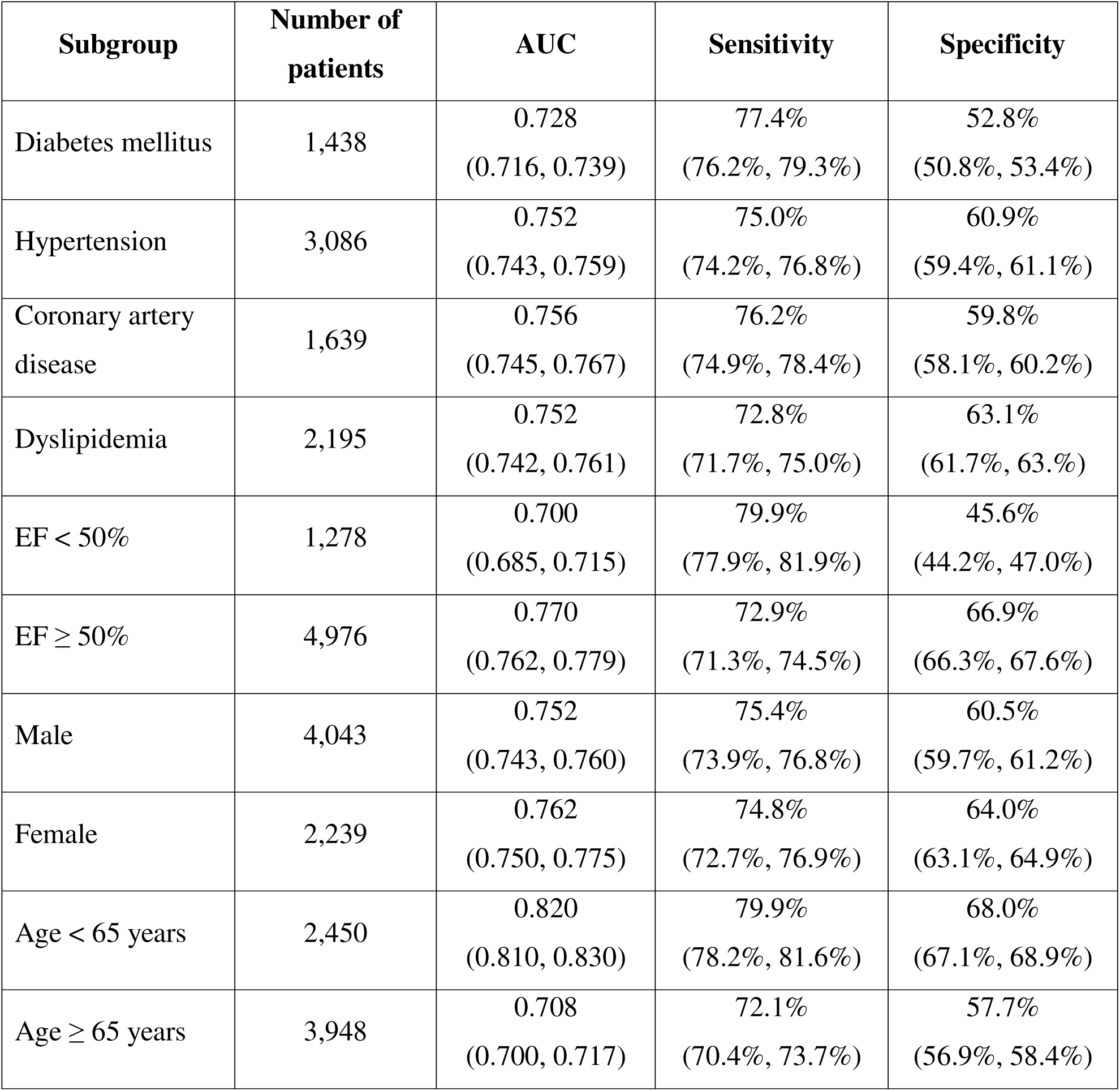
Performance of model in discriminating any stage of CKD by patient subgroup in the CSMC internal test cohort with 95% confidence intervals in parentheses.

During external validation, our model detected any stage of CKD in the KPNC cohort with AUC of 0.718 [0.714 – 0.723], sensitivity 77.2% [76.6% – 77.8%], and specificity 52.5% [52.1% – 52.9%] and in the SHC cohort with AUC of 0.719 [0.704 – 0.735], sensitivity 67.1% [64.7% – 69.6%], and specificity 65.1% [63.4% – 66.8%]. Performance in detecting mild, moderate, severe, and end-stage CKD in the KPNC external validation cohort had AUCs ranging from 0.678 to 0.838 and sensitivity from 62.4% to 88.1% (Supplementary 3). Model performance in the KPNC and SHC cohorts was similar across subgroups by sex, age, LVEF, hypertension, diabetes mellitus, dyslipidemia, and CAD (Tables 3 and 4).

**Table 3.** Performance of model in discriminating any stage of CKD by patient subgroup in the KPNC external validation cohort with 95% confidence intervals in parentheses.

| Subgroup | Number of patients | AUC | Sensitivity | Specificity |
| --- | --- | --- | --- | --- |
| Diabetes mellitus | 16,671 | 0.684<br>(0.678, 0.691) | 81.1%<br>(80.4%, 81.8%) | 40.8%<br>(40.0%, 41.5%) |
| Hypertension | 33,445 | 0.691<br>(0.687, 0.696) | 78.1%<br>(77.4%, 78.7%) | 46.4%<br>(45.9%, 46.9%) |
| Coronary artery disease | 20,399 | 0.698<br>(0.693, 0.704) | 80.9%<br>(80.2%, 81.6%) | 43.6%<br>(42.9%, 44.2%) |
| Dyslipidemia | 29,878 | 0.706<br>(0.701, 0.711) | 78.6%<br>(77.9%, 79.2%) | 48.1%<br>(47.6%, 48.7%) |
| EF < 50% | 1,915 | 0.659<br>(0.635, 0.675) | 86.4%<br>(83.6%, 87.7%) | 30.5%<br>(29.5%, 33.8%) |
| EF ≥ 50% | 8,431 | 0.716<br>(0.706, 0.729) | 74.4%<br>(72.5%, 76.0%) | 54.8%<br>(54.1%, 56.1%) |
| Male | 21,374 | 0.713<br>(0.707, 0.719) | 78.6%<br>(77.8%, 79.4%) | 50.0%<br>(49.3%, 50.6%) |
| Female | 20,184 | 0.725<br>(0.719, 0.732) | 75.8%<br>(74.8%, 76.8%) | 55.1%<br>(55.5%, 55.7%) |
| Age < 65 years | 11,947 | 0.786<br>(0.777, 0.794) | 77.8%<br>(76.5%, 79.2%) | 62.6%<br>(61.8%, 63.4%) |
| Age ≥ 65 years | 29,742 | 0.690<br>(0.684, 0.695) | 77.2%<br>(76.5%, 77.9%) | 47.9%<br>(47.4%, 48.5%) |

**Table 4.** Performance of model in discriminating any stage of CKD by patient subgroup in the SHC external validation cohort with 95% confidence intervals in parentheses.

| Subgroup | Number of patients | AUC | Sensitivity | Specificity |
| --- | --- | --- | --- | --- |
| Diabetes mellitus | 519 | 0.704<br>[0.669, 0.741] | 68.9%<br>[64.1%, 73.8%] | 63.6%<br>[59.2%, 67.9%] |
| Hypertension | 868 | 0.683<br>[0.654, 0.712] | 64.8%<br>[60.4%, 69.2%] | 61.1%<br>[57.9%, 64.4%] |
| Coronary artery disease | 894 | 0.638<br>[0.595, 0.680] | 63.1%<br>[57.5%, 68.8%] | 54.8%<br>[49.6%, 69.8%] |
| Dyslipidemia | 418 | 0.695<br>[0.667, 0.723] | 63.3%<br>[59.0%, 67.6%] | 63.8%<br>[60.7%, 66.9%] |
| EF < 50% | 469 | 0.683<br>[0.646, 0.720] | 79.2%<br>[75.0%, 83.3%] | 45.9%<br>[41.0%, 50.8%] |
| EF ≥ 50% | 1,756 | 0.695<br>[0.672, 0.718] | 60.8%<br>[57.1%, 64.6%] | 67.6%<br>[65.5%, 69.6%] |
| Male | 806 | 0.687<br>[0.656, 0.717] | 60.1%<br>[55.2%, 64.9%] | 66.2%<br>[62.9%, 69.3%] |
| Female | 950 | 0.695<br>[0.660, 0.730] | 61.9%<br>[55.8, 67.8%] | 68.6%<br>[66.0%, 71.2%] |
| Age < 65 years | 872 | 0.745<br>[0.715, 0.776] | 62.4%<br>[56.6%, 68.1%] | 74.3%<br>[71.6%, 76.9%] |
| Age ≥ 65 years | 885 | 0.641<br>[0.608, 0.673] | 59.6%<br>[54.6%, 64.7%] | 60.6%<br>[57.5%, 63.6%] |

### Error Mode Analysis

We analyzed patients whom the AI model identified patients as having CKD, but who did not have a corresponding ICD-10 code. In the CSMC internal test cohort, 2,778 (44.2%) of patients were identified as having CKD based on their PLAX video without a clinical diagnosis. Of these patients, 2,541 patients had eGFR measured within 1 week of echocardiography, and 2,182 (85.9%) patients had had measured eGFR < 90 mL/min/1.73 m^2^ within 1 week of echocardiography. 1,202 patients had longitudinal eGFR values, with 657 (54.7%) patients having a history of at least two measured eGFR < 90 mL/min/1.73 m^2^ at least 90 days apart; 276 (42.0%) patients had eGFR < 60 mL/min/1.73 m^2^, corresponding to Stage 3a CKD or worse. Using their eGFR and UACR measurements, we found that 111 (4.0%) patients had KDIGO risk scores corresponding with moderately to very high increased risk.

Similarly, in the KPNC external validation cohort, 16,019 (38.5%) patients were identified by the model as having CKD but did not have a clinical diagnosis. 7,319 (45.7%) patients had eGFR measured within 1 week of their study. All 7,319 patients had eGFR < 90 mL/min/1.73 m^2^, and eGFR < 60 mL/min/17.3 m^2^ was measured in 1,057 (14.4%) patients. 1,737 patients had longitudinal creatinine values measured, with 113 (6.5%) patients had at least two measured eGFR < 90 mL/min/1.73 m^2^ at least 90 days apart. Additionally, 426 (2.7%) patients had albuminuria with UACR ≥ 30 within one month of their PLAX video. KDIGO risk scores could be calculated for 557 (3.5%) patients, with 251 (45.1%) patients having moderate to very high risk KDIGO scores. In the SHC cohort, 773 patients were identified by the model as having CKD although they did not have a clinical diagnosis. 365 (47.2%) patients had eGFR < 90 mL/min/1.73 m^2^ within 1 week of echocardiography, and 79 (10.2%) patients had eGFR < 60 mL/min/1.73 m^2^. Of the 773 false-positive patients, 365 patients had longitudinal creatinine values. 181 (49.6%) patients had at least two measured eGFR values < 90 mL/min/1.73 m^2^ at least 90 days apart, and 106 (29.0%) patients with at least two measured eGFR < 60 mL/min/1.73 m^2^ at least 90 days apart. UACR values were not available for patients at SHC, and KDIGO risk scores were not calculated.

### Model Interpretability

To support explainability for clinical use, we generated GradCAM heatmaps of significant pixels for CKD detection. GradCAM highlighted the mitral valve and left atrium as important features for AI model decision-making (Figure 2). These results are consistent with existing literature highlighting excess prevalence of mitral valve dysfunction that occurs in the early stages of CKD, with patients with CKD having higher rates of mitral regurgitation and undergoing mitral valve calcification 10-20 years earlier than those without CKD^38–40^.

**Figure 2.**
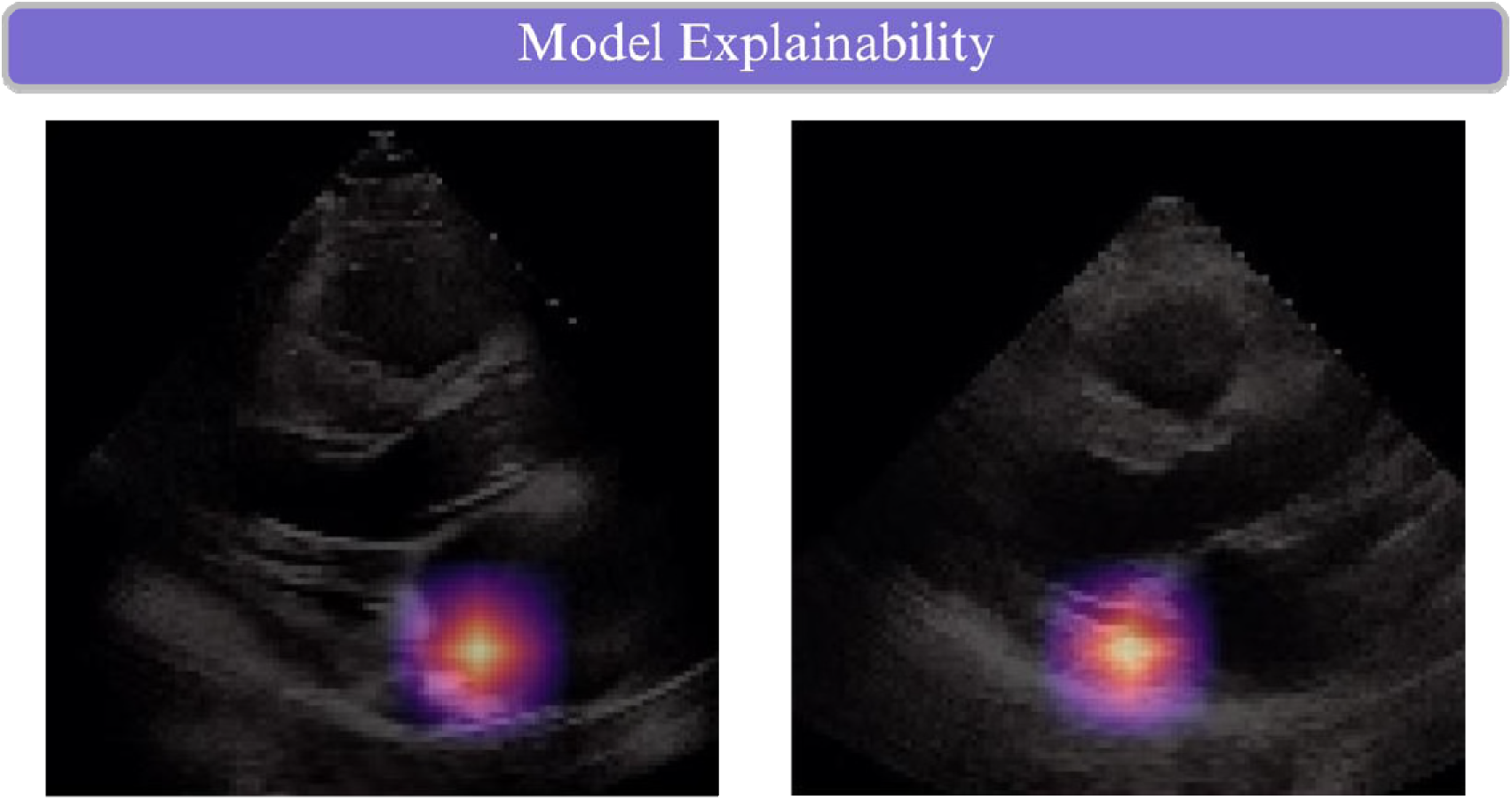
Grad-CAM^37^ activation was primarily at the mitral valve in PLAX videos.

## Discussion

In this work, we developed a deep learning model to detect chronic kidney disease (CKD) from parasternal long axis (PLAX) echocardiogram videos. Our model demonstrated strong performance to identify any stage of CKD across diverse patient subgroups. We validated our model in two geographically distinct centers, emphasizing its ability to generalize across different patient populations and clinical protocols. With an overall sensitivity of 76%, our study offers an innovative avenue to screen for CKD. Improved detection can connect patients to treatment earlier in their disease course and decrease the growing burden of CKD.

CKD affects 850 million individuals globally and is projected to be the 5^th^ leading cause of lost years of life by 2040^1^. The rising prevalence of CKD reflects a rise in its metabolic risk factors of hypertension, diabetes, and obesity^1,15^. These factors also increase risk of CVD, with renal dysfunction contributing additive risk^7,14,15^. This interplay between metabolic, renal, and cardiovascular disease is captured by cardiovascular-kidney-metabolic (CKM) syndrome, which in turn increases morbidity and premature mortality. Early recognition of CKD and CKM can connect patients to appropriate management and slow disease progression. There is a critical need to improve identification of CKD syndrome. However, current methods to detect CKD have low screening rates.

Existing ML studies predict risk of CKD using demographic, clinical, and laboratory values^31–35^. While these models show promise for detection, they require serum values or physical exam findings. Other models predict progression to ESRD in patients with established CKD, rather than detecting new-onset CKD^34,35^. Holmstrom, et al. utilized 12-lead electrocardiograms to detect CKD within a 1-year window^36^. Identifying CKD within a narrower window can improve disease detection and facilitate timely treatment to prevent CKD progression.

Our model offers a noninvasive, rapid avenue to detect CKD from PLAX videos with high fidelity. Performance was consistently strong in high-risk patients, including those with diabetes, hypertension, and dyslipidemia (Tables 2, 3). Because our ML-based screening analyzes echocardiography, it may be potentially easier to use in a clinical setting, whereas the necessary variables for other calculators may not always be obtained from health records. Moreover, our work detects any stage of CKD to facilitate earlier detection, which can complement existing studies that focus on progression to ERSD^34,35,44^.With sensitivities from 72.1% to 81.1%, our model can be leveraged for initial screening of occult or early-stage CKD. Combined with clinical histories, our ML algorithm may address diagnostic gaps for high-risk patients, for whom screening is not regularly performed^6,9^. Our model also identified patients without a clinical diagnosis of CKD, but based on their eGFR, UACR, and KDIGO risk score, may have subclinical or early-stage CKD and may benefit from intervention. With the American Heart Association’s recent recommendations to screen for and stage CKM syndrome, we envision that our deep learning model can be integrated into clinical workflows for simultaneous evaluation of cardiac and renal function^15^. Early identification of CKD can enable earlier intervention, improve patient awareness, and slow disease progression.

Explainability analysis highlights that our model may be learning imaging biomarkers indicative of disease pathophysiology. GradCAM findings revealed that our algorithm focused on the mitral valve and left atrium. Dysfunction of these two structures are early manifestations of CKD-induced cardiac disease. Moreover, our model performed consistently across subgroups, suggesting that it did not rely on co-morbidities for its predictions. Rather, our model likely learned disease features shared amongst high-risk patients.

Future work involves validation of our model at additional sites and integration of our model into clinical workflows to characterize its clinical utility. Given the need for CKD screening in high-risk patient populations, we anticipate that ML-based detection can increase detection rates, facilitate early intervention for patients, and help decrease the global disease burden of CKD.

In conclusion, we developed and externally validated a deep learning model to detect CKD from PLAX echocardiography videos. Our algorithm can detect any stage of CKD, including early stages that may go undiagnosed, suggesting deep learning screening for CKD from echocardiography can help bolster low detection rates and promote more timely management for physicians.

## Methods

### Data sources and study population

To develop our deep learning model, we retrospectively analyzed a cohort of patients who received care at Cedars-Sinai Medical Center (CSMC) between 2016 and 2022. Similarly, we identified two patient cohorts for external validation – one at Kaiser-Permanente Northern California (KPNC) with patients who received care between 2022 and 2025 and a second at Stanford Healthcare (SHC) with patients who received care between 2010 and 2018. In both the development and external validation cohorts, the case cohort consisted of patients who were diagnosed with CKD by International Classification of Disease (ICD)-10 code (Supplementary Table 1) and who had a PLAX echocardiogram within 1 month. Control patients were matched to cases by age and sex in a 1:5 ratio. For patients with a creatinine measurement within one week of echocardiogram acquisition, estimated glomerular filtration rate (eGFR) was calculated. Urine albumin and creatinine labs within one month of echocardiogram acquisition were extracted to calculate UACR. Demographic and clinical co-morbidities were extracted from the electronic health records using ICD-10 codes. Echocardiographic parameters of left ventricular ejection fraction (LVEF) and LV volumes were obtained from the final study reports.

### Model Development and Interpretability

We trained an R(2+1)D convolutional neural network as a binary classifier to predict the presence of CKD within 1 month of PLAX echocardiogram acquisition. The dataset was split into train, validation, and test sets in an 8:1:1 ratio on the patient-level. If a patient had multiple PLAX videos, each video was treated as an independent sample. We then trained our model with a learning rate of 1e-3, Adam optimization, and binary cross-entropy loss. Early stopping was implemented based on validation loss. Model interpretability was analyzed with Gradient-weight Class Activation Mapping (Grad-CAM)^37^ to generate heatmaps of pixel importance for predictions.

### Statistical Analysis

Model performance was assessed using area under the receiver operator characteristic curve (AUC), sensitivity, and specificity on the test set, which was not seen during model development and training. The 95% confidence interval (CI) was calculated with 10,000 bootstrapped samples. We evaluated performance to detect mild, moderate, severe, and end-stage CKD as defined by ICD-10 codes. The ability of the model to discriminate patients with documented CKD and laboratory values for eGFR or albuminuria was also assessed. Subgroup analysis was performed based on co-morbidities of diabetes mellitus, hypertension, dyslipidemia, and coronary artery disease, and myocardial disease; demographics of sex and age greater or lower than 65 years old; and cardiac function based on LVEF.

### Error Mode Analysis

We identified patients who were classified as having CKD by the model but did not have a corresponding ICD-10 code, and patients who had an ICD-10 code for CKD but were not identified by the model. We analyzed their UACR performed within 31 days of echocardiography. eGFR measurements for patients over 3 months to 1 year were obtained and compared against diagnostic criteria for CKD. Finally, the Kidney Disease: Improving Global Outcomes (KDIGO) risk score was also calculated.

## Data and Code Availability

Model weights and code are publicly available on Github at https://github.com/echonet/ckd. Due to its potentially identifiable nature, the imaging dataset will not be released.

---

**Supplementary Figure 1.**
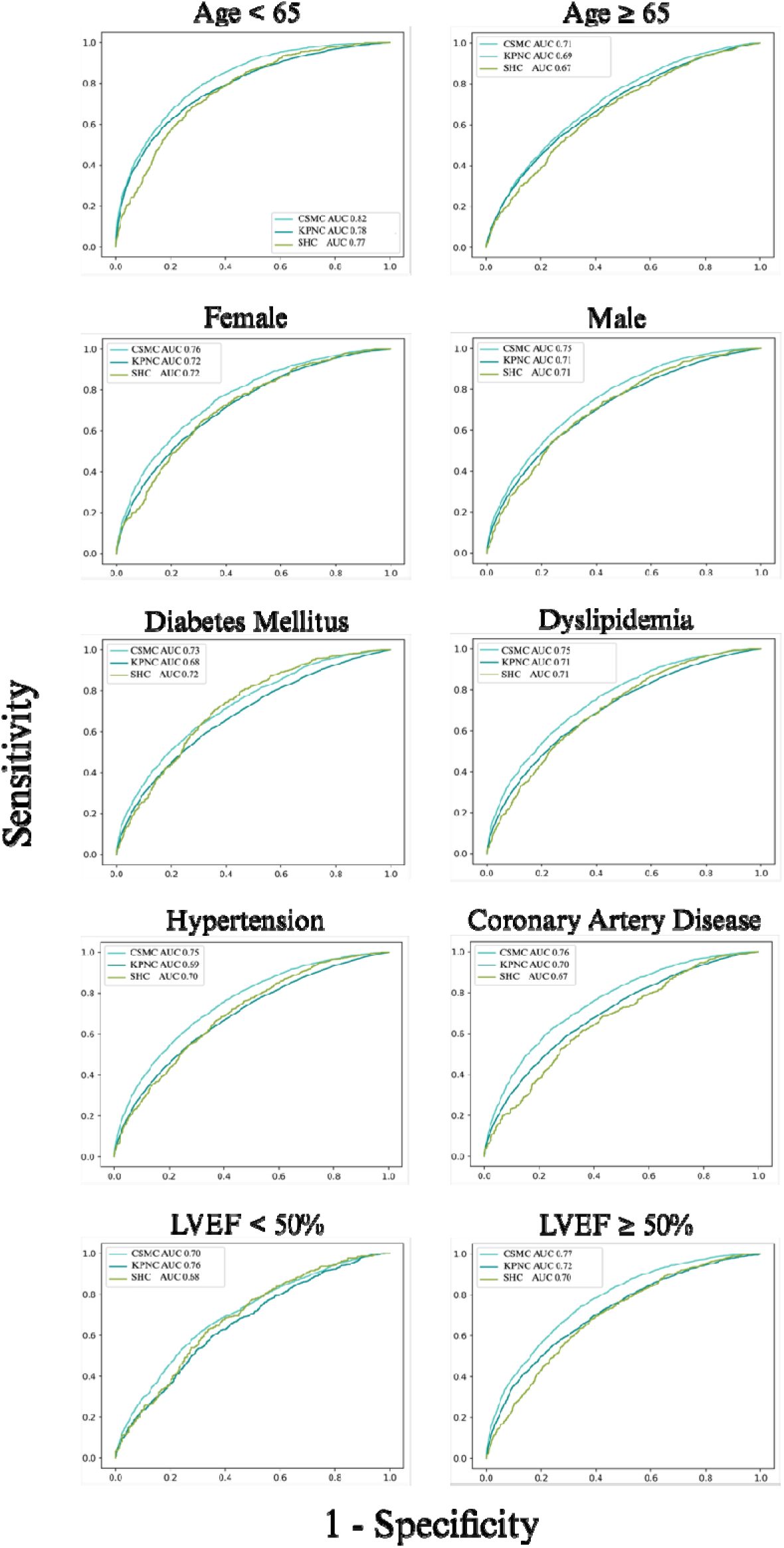
Model performance in patient subgroups based on diabetes mellitus, hypertension, dyslipidemia, coronary artery disease, ejection fraction, age, and sex in the CSMC internal test and KPNC external validation cohorts.

**Supplementary Table 1.** ICD-10 codes used to identify patients diagnosed with CKD.

|  |  |
| --- | --- |
| **ICD 10 Codes** | N18.1 |
|  | N18.2 |
|  | N18.30, N18.31, N18.32 |
|  | N18.4 |
|  | N18.5 |
|  | N18.6 |
|  | N18.9 |

**Supplementary Table 2.** Demographics and co-morbidities of the patients in the train, validation, and test cohorts at CSMC for model development.

|  | **Train**<br>**(n=50,254)** | **Validation**<br>**(n=6,282)** | **Test**<br>**(n=6,282)** |
| --- | --- | --- | --- |
| Age | 67.3 ± 15.7 | 66.8 ± 16.0 | 67.0 ± 15.2 |
| Sex |  |  |  |
| Male | 32,312 (64.3%) | 4,021 (64.0%) | 4,043 (64.4%) |
| Female | 17,942 (35.7%) | 2,261 (36.0%) | 2,239 (35.6%) |
| Race and Ethnicity |  |  |  |
| American Indian or Alaska Native | 109 (0.2%) | 10 (0.2%) | 15 (0.2%) |
| Asian | 3,562 (7.1%) | 448 (7.1%) | 479 (7.6%) |
| Black or African American | 6,663 (13.3%) | 820 (13.1%) | 827 (13.2%) |
| Native Hawaiian/Pacific Islander | 144 (0.3%) | 17 (0.3%) | 27 (0.4%) |
| Hispanic | 5,948 (11.8%) | 721 (1.5%) | 784 (12.5%) |
| Non-Hispanic | 42,430 (84.4%) | 5,348 (85.1%) | 5,264 (83.8%) |
| White | 34,518 (68.7%) | 4,358 (69.4%) | 4,220 (67.2%) |
| Other | 3,464 (6.9%) | 418 (6.7%) | 484 (7.7%) |
| Unknown | 1,258 (2.5%) | 123 (2.0%) | 131 (2.1%) |
| Left ventricular ejection fraction % | 56.3% ± 15.4% | 56.7% ± 15.2% | 56.2% ± 15.5% |
| Left ventricular end-diastolic volume index (mL/m<sup>2</sup>) | 47.5 ± 26.7 | 46.8 ± 23.1 | 47.6 ± 25.1 |
| Left ventricular end-systolic volume index (mL/m<sup>2</sup>) | 22.1 ± 17.8 | 22.0 ± 18.6 | 22.3 ± 17.3 |
| Estimated glomerular filtration rate (eGFR) | 66.3 ± 30.4 | 66.1 ± 31.2 | 65.6 ± 30.7 |
| Urine albumin-to-creatinine ratio (UACR) | 137.0 ± 309.5 | 159.6 ± 337.3 | 165.4 ± 354.2 |
| Coronary artery disease | 13,015 (25.9%) | 1,676 (26.7%) | 1,639 (26.1%) |
| Hypertension | 24,736 (49.2%) | 3,084 (49.1%) | 3,086 (49.1%) |
| Diabetes mellitus | 11,290 (22.5%) | 1,419 (22.6%) | 1,438 (22.9%) |
| Dyslipidemia | 17,503 (34.8%) | 2,212 (35.2%) | 2,195 (34.9%) |

**Supplementary Table 3.** Model performance in discriminating different degrees of CKD in the CSMC cohort (top) and KPNC cohort (bottom). Negative examples were defined as PLAX videos without CKD. 95% confidence intervals are presented in parentheses.

| **Cedars-Sinai Internal Test Cohort** |  |  |  |  |  |
| --- | --- | --- | --- | --- | --- |
| **Degree of CKD** | **Number of patients** | **ICD-10 Codes** | **AUC** | **Sensitivity** | **Specificity** |
| Mild | 93 | N18.1,<br>N18.2 | 0.694<br>(0.670, 0.718) | 68.6%<br>(63.4%, 73.2%) | 61.3%<br>(61.3%, 62.4%) |
| Moderate | 362 | N18.3,<br>N18.30,<br>N18.31 | 0.701<br>(0.689, 0.712) | 68.4%<br>(65.5%, 70.1%) | 61.3%<br>(61.3%, 62.4%) |
| Severe | 149 | N18.4,<br>N18.5 | 0.773<br>(0.755, 0.790) | 76.6%<br>(73.1%, 79.7%) | 61.3%<br>(61.3%, 62.4%) |
| ESRD | 307 | N18.6 | 0.840<br>(0.829, 0.850) | 86.9%<br>(84.6%, 88.3%) | 61.3%<br>(61.3%, 62.4%) |
| **Kaiser-Permanente External Validation Cohort** |  |  |  |  |  |
| **Degree of CKD** | **Number of patients** | **ICD-10 Codes** | **AUC** | **Sensitivity** | **Specificity** |
| Mild | 682 | N18.1,<br>N18.2 | 0.678<br>(0.661, 0.693) | 72.2%<br>(69.5%, 74.8%) | 52.5%<br>(52.0%, 52.9%) |
| Moderate | 5,832 | N18.3,<br>N18.30,<br>N18.31 | 0.679<br>(0.673, 0.684) | 72.8%<br>(71.9%, 73.7%) | 52.5%<br>(52.0%, 52.9%) |
| Severe | 1,088 | N18.4,<br>N18.5 | 0.785<br>(0.773, 0.796) | 84.8%<br>(83.0%, 86.4%) | 52.5%<br>(52.0%, 52.9%) |
| ESRD | 1,360 | N18.6 | 0.830<br>(0.821, 0.839) | 88.1%<br>(86.8%, 89.4%) | 52.5%<br>(52.0%, 52.9%) |
| **Stanford Healthcare External Validation Cohort** |  |  |  |  |  |
| **Degree of CKD** | **Number of patients** | **ICD-10 Codes** | **AUC** | **Sensitivity** | **Specificity** |
| Mild | 337 | N/A | 0.686<br>(.665, .707) | 62.4%<br>(58.9%, 65.8%) | 65.1%<br>(63.4%, 66.8%) |
| Moderate | 420 | N/A | 0.742<br>(.727, .757) | 70.3%<br>(67.8%, 72.6%) | 65.1%<br>(63.4%, 66.7%) |
| Severe | 125 | N/A | 0.838<br>(.819, .857) | 85.4%<br>(81.7%, 89.0%) | 65.1%<br>(63.4%, 66.7%) |
| ESRD | 129 | N/A | 0.837<br>(.818, .855) | 82.3%<br>(78.8%, 85.7%) | 65.1%<br>(63.4%, 66.8%) |

